# Association between BMI and cause-specific long-term mortality in acute myocardial infarction patients

**DOI:** 10.1101/2024.05.15.24307457

**Authors:** Timo Schmitz, Dennis Freuer, Philip Raake, Jakob Linseisen, Christa Meisinger

**Affiliations:** Epidemiology, Medical Faculty, University of Augsburg, Augsburg, Germany; University Hospital Augsburg, Department of Cardiology, Respiratory Medicine and Intensive Care, Augsburg, Germany

**Keywords:** BMI, acute myocardial infarction, long-term mortality, cause-specific mortality

## Abstract

**Background:** To investigate the association between body mass index (BMI) at acute myocardial infarction (AMI) and all-cause as well as cause-specific long-term mortality.

**Methods:** The analysis was based on 10,651 hospitalized AMI patients (age 25-84 years) recorded by the population-based Myocardial Infarction Registry Augsburg between 2000 and 2017. The median follow-up time was 6.7 years [IQR: 3.5-10.0)]. Cause-specific mortality was obtained by evaluating the death certificates. In multivariable-adjusted COX regression models using cubic splines for the variable BMI, the association between BMI and cause-specific mortality (all-cause, cardiovascular, ischemic heart diseases, cancer) was investigated. Additionally, a subgroup analysis in three age groups was performed for all-cause mortality.

**Results:** Overall, there was a statistically significant U-shaped association between BMI at AMI and long-term mortality with the lowest hazard ratios (HR) found for BMI values between 25 and 30 kg/m². For cancer mortality, higher BMI values > 30 kg/m² were not associated with higher mortality. In patients aged <60 years, there was a significant association between BMI values > 35 kg/m² and increased all-cause mortality; this association was missing in 60 to 84 years old patients. For all groups and for each specific cause of mortality, lower BMI (< 25kg/m²) values were significantly associated with higher mortality.

**Conclusions:** Overall, a lower BMI – and also a high BMI in patients younger than 60 years - seem to be a risk factors for increased all-cause mortality after AMI. A BMI in a mid-range between 25 and 30 kg/m² is favorable in terms of long-term survival after AMI.

**What is Known?:**

- Obesity is considered a major risk factor for cardiovascular diseases and coronary artery disease.
- Nonetheless, the results of prior studies indicated lower mortality for overweight and obese individuals after acute myocardial infarction (AMI); a phenomenon often called ‘obesity paradox’.

**What the Study Adds:**

- This study analyzed the association between body mass index (BMI) and cause-specific long-term mortality after AMI in a population-based cohort of 10.000 AMI patients recorded by the Myocardial Infarction Registry Augsburg.
- The obesity paradox was partially confirmed, as we found a U-shaped association between BMI and long-term mortality with the lowest risk of mortality in overweight individuals.
- For cancer mortality and in elderly, obesity (BMI > 30 kg/m²) was not associated with mortality. Lower BMI < 25 kg/m² went along with significantly higher mortality in all age groups and for all causes of death (all-cause, cardiovascular, ischemic heart disease, cancer)

## 1. Introduction

Extreme body mass indices (BMI) are known to be a major risk factor for several diseases. Very low BMI (e.g. in anorexia or cancer cachexia) as well as very high values in obese individuals go along with increased risk of mortality ^1–3^. Especially obesity is considered to be an important risk factor for metabolic diseases like diabetes mellitus and, closely linked to that, a risk factor for several cardiovascular diseases as well ^4–6^. However, for patients with coronary artery disease (CAD) or acute myocardial infarction, prior studies have reported conflicting results regarding the association between high BMI values and long-term mortality: Some researchers found decreased mortality for patients with obesity, often referred to as the ‘obesity paradox’ ^7–14^; a phenomenon that was questioned by the results of several other studies ^15,16^. Thus, the true nature of the association between BMI and long-term mortality in AMI survivors remains controversial. The present study aims to contribute to a clearer and deeper understanding of this complex relationship. To do so, a large unselected sample with high quality data is required, which in this case is derived from the population-based Myocardial Infarction Registry Augsburg. Next to BMI and all-cause mortality, there is also a lack of knowledge on the association between BMI and cause-specific mortality, like cardiovascular diseases (CVD) mortality, ischemic heart disease mortality or cancer mortality. Finally, the impact of age on the associations between BMI and mortality will be investigated.

## 2. Methods

### 2.1 Study population

The study used data from the Myocardial Infarction Registry in Augsburg. It was initially established in 1984 as part of the World Health Organization (WHO) project MONICA (Monitoring Trends and Determinants in Cardiovascular Disease). For almost 40 years, all cases of non-fatal AMI and coronary deaths in the study region (city of Augsburg and the two adjacent counties, approximately 680,000 inhabitants) were registered. Inclusion criteria for patients were: primary residence in the study region and age between 25 and 74 years (until 2008) and 25-84 years (from 2009-2017), respectively.

Detailed information on case identification, classification and quality control of the data can be found in a previous publication ^17^. All study participants gave written informed consent. Methods of data collection and questionnaires have been approved by the ethics committee of the Bavarian Medical Association (Bayerische Landesärztekammer) and the study was performed in accordance with the Declaration of Helsinki. The study was registered at the German Register of Clinical Studies (DRKS, project number DRKS00029042).

### 2.2 Data collection

During their hospital stays, patients were interviewed by trained study nurses using a standardized questionnaire. Additionally, the patients’ medical chart was reviewed in detail to confirm the information obtained by the interview and to collect additional data. By this procedure, a wide range of demographic data, data on cardiovascular risk factors, medical history, comorbidities, laboratory values, diagnostics, treatment and medication were collected for each patient.

In particular, information about body weight and height were obtained from the patient’s medical file. The variable BMI was calculated as weight in kg divided by the squared height in meters. Renal function according to the estimated glomerular filtration rate (eGFR) was calculated using the CKD-EPI formula.

### 2.3 Outcome

The outcome of this study was all-cause and cause-specific long-term mortality. Mortality was ascertained by regularly checking the vital status of all registered AMI patients in cooperation with the regional population registries. Death certificates were obtained from local health departments. Additionally, for each deceased patient, a standardized questionnaire was sent out to the former general practitioner and/or coroner. The questionnaire includes, among other topics, questions about preexisting comorbidities and diseases, circumstances of the acute event (e.g., reanimation) and specific medication. The main cause of death, encoded by the ICD-10 classification ^18,19^, was determined on the base of the death certificate. CVD deaths included all deaths encoded by I00 to I99, ischemic heart diseases were defined as deaths encoded by I20 to I25 and finally, cancer deaths included the ICD-10 codes C00 to D48.

### 2.4 Statistical analysis

For categorical variables, the results were presented as absolute frequencies with percentages and the Chi-square test was used to test for differences between groups. For normally-distributed continuous variables, the results were presented as means with standard deviations and we used Student’s t-tests to test for differences. For non-normally distributed continuous variables the results were presented as medians with inter-quartiles ranges and nonparametric tests were applied.

As there was no follow-up information on BMI over time, we limited the post-AMI observational time to 10 years; patients with longer observational times were censored after 10 years. To concentrate on long-term mortality, all patients with an observational time of 28 days and less (the common definition of short-term mortality in the Augsburg Myocardial Infarction Registry is death within 28 days after AMI) were excluded from the analysis.

### 2.5 COX regression analyses

Multivariable-adjusted COX proportional hazard regression models were used to analyze the association between BMI and all-cause and cause-specific long-term mortality (CVD mortality, ischemic heart disease (IHD) mortality and cancer mortality). The relationship between BMI and mortality was modeled using restricted cubic splines (RCS) with 4 knots (at the 5th, 35th, 65th and 95th percentiles), which allows to capture a potential non-linear association; the reference BMI value was set to 27.5 kg/m². The models were adjusted for the continuous variable age (in years) and the categorical variables sex (male/female), diabetes (yes, no), hypertension (yes, no), smoking (current, ex-smoker, never smoker, smoking status unknown), renal function according to eGFR (≥ 60ml/min, 30-59ml/min, < 30ml/min, unknown), left-ventricular ejection fraction (EF) (≤30%, >30%, unknown), type of infarction (STEMI, NSTEMI, other/unknown), percutaneous coronary intervention (PCI) (yes, no) and bypass therapy (yes, no).

The proportional hazard assumptions were checked graphically by inspecting the Kaplan Meier survival curves as well as log(-log(survival)) plots; for the continuous variables BMI and age the survival curves stratified for the corresponding quartiles were inspected.

In a subgroup analysis, we calculated the COX regression models stratified for three age groups: age < 60 years between 60 and 74 years, and ≥ 74 years.

Furthermore, we calculated all models not using splines, but quartiles for the variable BMI (displayed in the supplementary material). Finally, we performed two sensitivity analyses: First, we calculated the COX model for the outcome ‘cancer mortality’ only including cases with a follow-up time of two years and more (also excluding deaths within the first 2 years). Second, we calculated all models only for patients who never smoked cigarettes (also displayed in the supplementary material).

All statistical analyses were carried out with the R program version 4.3.2. A p value of < 0.05 was considered as statistically significant.

## 3. Results

Baseline characteristics of the study sample were displayed in table 1. The mean age was 63.7 (11.1) years and 73.3% of all patients were men. During the observational time of up to ten years, 2,770 deaths were recorded among the 10,651 AMI patients. In total, 1,435 (51.8%) deaths were classified as CVD deaths, 1,042 (37.6%) deaths were due to ischemic heart disease and 577 (20.8%) due to cancer. Elderly patients ≥75 years were less likely to die because of cancer and had more comorbidities (diabetes, hypertension, impaired renal function) compared to AMI patients < 75 years; the percentage of current smokers decreased from 58.0% among patients < 60 years to only 7.0% in the group ≥ 75 years. PCI was performed in 70.5% of all cases, with the highest frequency in the age group < 60 years and the lowest frequency among the elderly, aged ≥ 75 years.

**Table 1:**
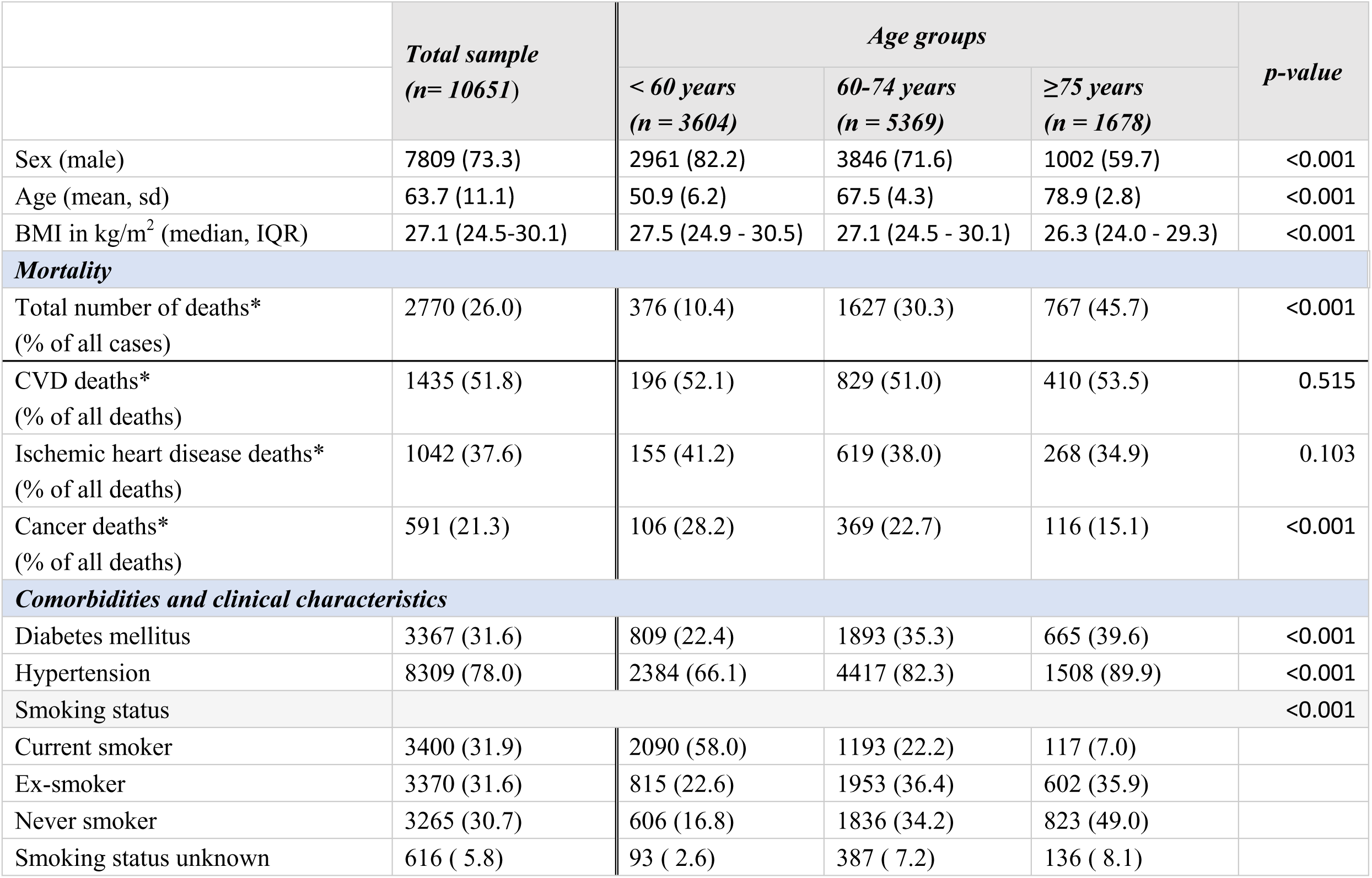
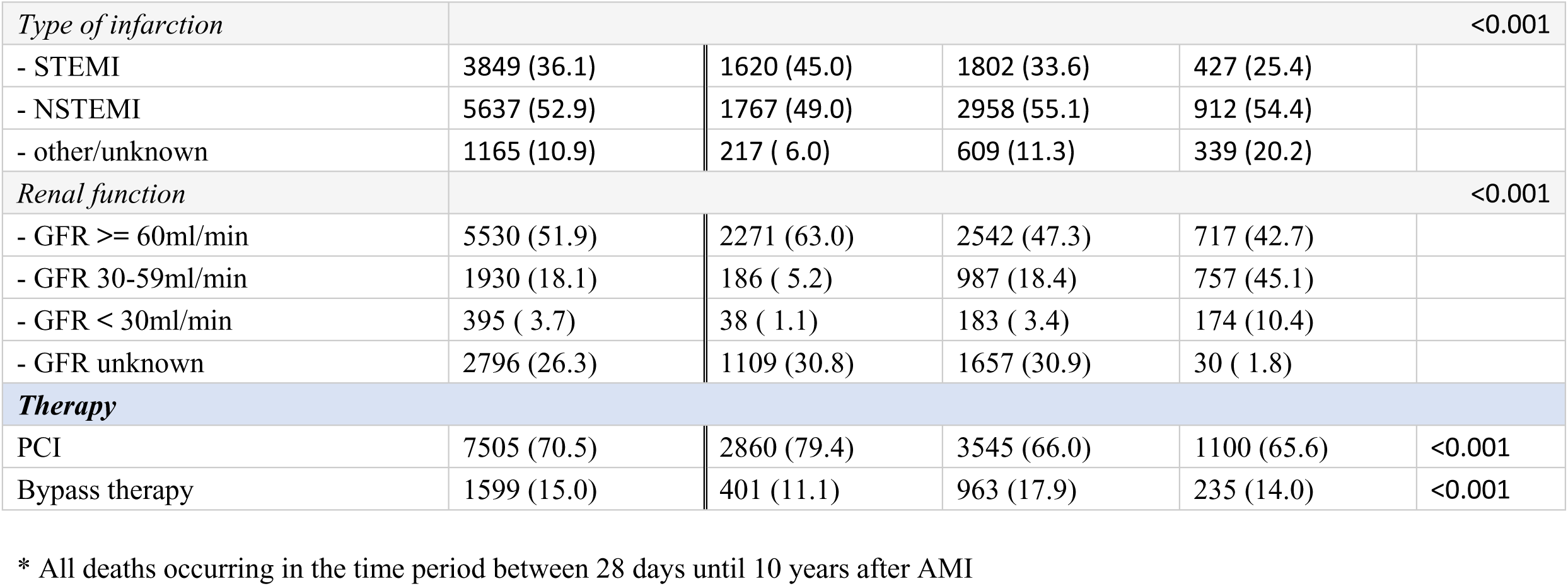
Baseline characteristics of patients with and without diabetes. Categorical data presented as total numbers (%). Numeric data is presented as mean (SD) or median (IQR).

### 3.1 All-cause and cause-specific mortality - total sample

There was a U-shaped association between BMI and long-term all-cause mortality in the total sample (figure 1A). Compared to the reference BMI value of 27.5 kg/m², patients with a BMI of 20 kg/m² had a highly increased risk of mortality (HR 2.14 [1.91,2.40]); patients with BMI values of 25 and 30 kg/m² had HRs of 1.04 [0.99,1.10] and 1.08 [1.03,1.14] respectively. Obese patients again had an increased risk of mortality with HRs of 1.22 [1.12,1.33] and 1.36 [1.15,1.61]. Table S1 displays HR values and 95%CI for specific BMI values and for each outcome). The U-shaped relation for all-cause mortality was confirmed by the COX model using BMI quartiles: the first and the fourth quartile had significantly higher HR values compared to the second quartile (reference group) (supplementary table S2). A similar result was obtained for CVD mortality (figure 1B). For ischemic heart diseases there appeared to be a U-shaped association as well (figure 1C), but the results were no longer significant for high BMI values. A different constellation was found for cancer mortality: low BMI levels were still strongly associated with higher mortality, but for patients with high BMI values we didńt observe a higher cancer mortality risk (figure 1D).

**Figure 1:**
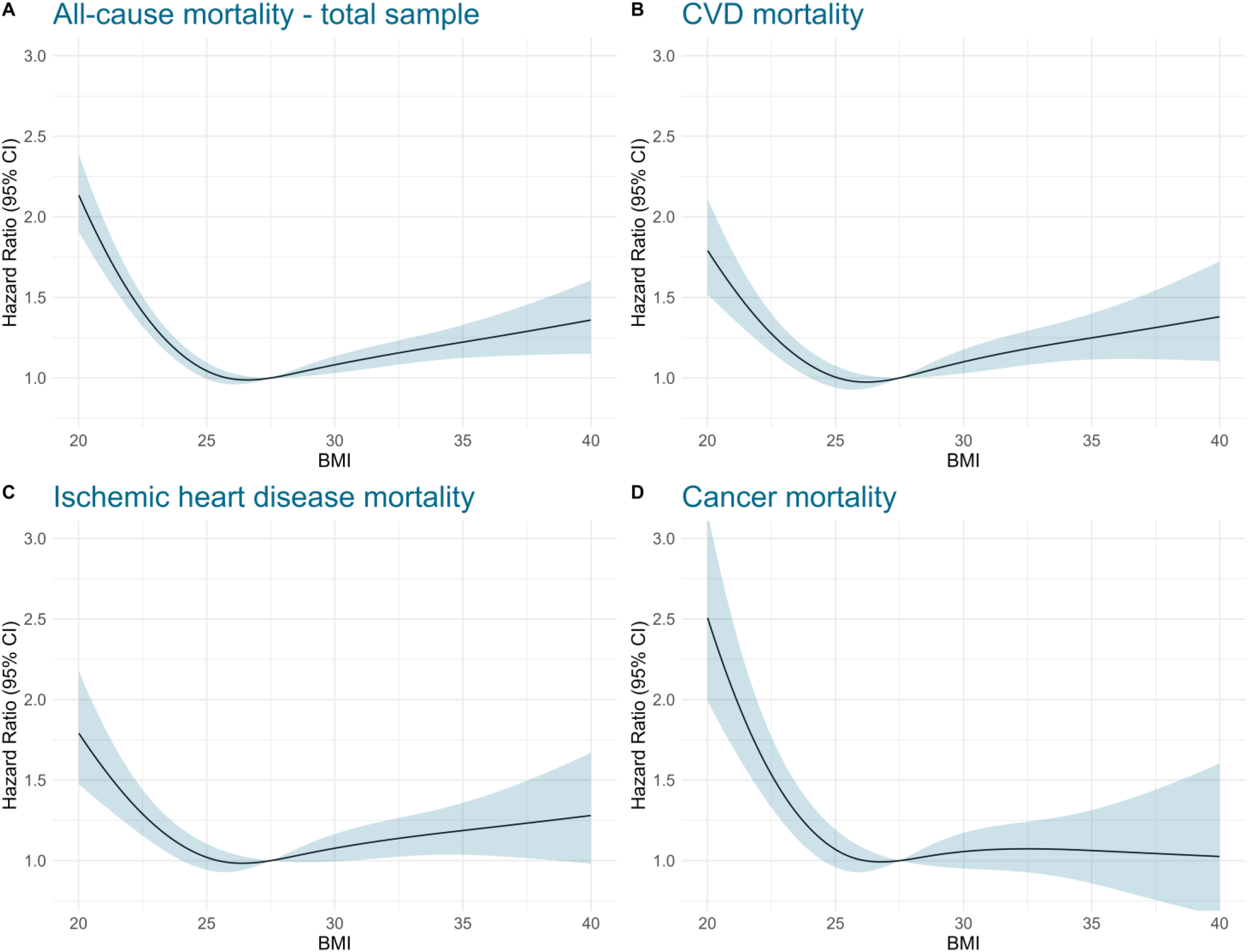
Association between BMI and all-cause and cause specific mortality in the total sample. The COX regression models were adjusted for age, sex, diabetes, hypertension, smoking status, eGFR, left-ventricular EF, type of infarction, PCI and bypass therapy.

### 3.2 All-cause mortality in different age groups

For the age group < 60 years, there was a pronounced U-shaped association between BMI and all-cause mortality (figure 2A). Nevertheless, in the regression model using BMI quartiles, only the first, but not the fourth BMI quartile was significantly associated with mortality compared to the second quartile (reference), see table S2. In the age group 60-74 years there was also a U-shaped association (figure 2B) and a significant higher mortality for patients in the first and fourth quartile, see table S2. For the elderly group ≥ 75 years however, higher BMI levels were not associated with higher mortality (figure 2C and table S2). Hence, low BMI levels were significantly associated with an elevated risk of dying for all age groups, (figure 2), but only for AMI patients < 75 years high BMI values were also significantly associated with increased mortality.

**Figure 2:**
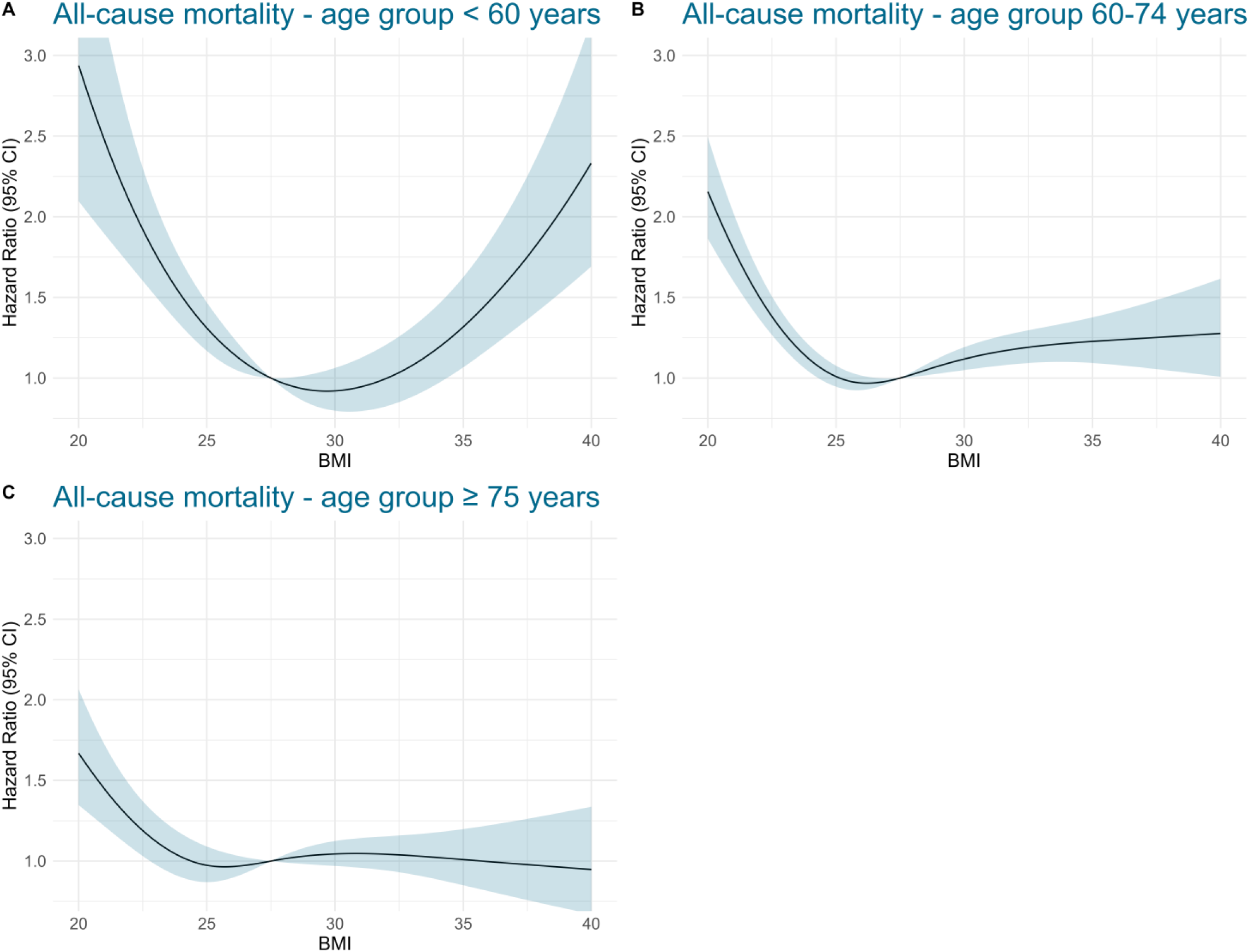
Association between BMI and all-cause mortality in different age groups. The COX regression models were adjusted for age, sex, diabetes, hypertension, smoking status, eGFR, left-ventricular EF, type of infarction, PCI and bypass therapy.

### 3.3 Sensitivity analyses

Excluding all cases with an observational time of less than two years had only minor impact on the association between BMI and cancer mortality, see table S2 and figure S1 (supplementary material). Moreover, including only never smokers into the COX models had moderate effect on the results: While the general shape of the associations between BMI and all-cause and cause-specific mortality was confirmed (in particular lowest mortality for BMI between 25 and 30 kg/m^2^), the strengths of the associations appeared to be moderately attenuated (in terms of lower HR values compared to the main models), see figures S2 and S2, supplementary material.

## 4. Discussion

In this study, we found an overall U-shaped association between BMI and long-term mortality after AMI. For the older age group (>75 years) as well as for cancer mortality, however, high BMI values were not associated with higher risk for early mortality.

### High BMI values and long-term mortality

Several prior studies have examined the relationship between (high) BMI and long-term mortality after myocardial infarction, and thereby reporting conflicting results. Some studies and meta-analyses indicated a survival benefit for patients with higher BMI in CAD, acute coronary syndrome and AMI ^7–14^; a phenomenon often referred to as the ‘obesity paradox’. This phenomenon is only partially confirmed by the present study, as we found the lowest mortality for patients with BMI values between 25 and 30 kg/m², which is categorized as ‘pre-obesity’ or ‘overweight’ by the WHO classification ^20^. For BMI > 30 kg/m², which is classified as obesity, mortality began to increase. These observations are supported by a recent study from Sweden analyzing over 25,000 STEMI patients; they found the lowest 1-year all-cause mortality for the overweight group (BMI 25.0–29.9 kg/m^2^) ^21^.

In fact, there are prior studies and meta-analyses that question this ‘obesity paradox’ in CAD and AMI patients ^15,16^. One example is a recent study by Al-Shaar et al. ^16^; they found higher mortality for patients with higher BMI for all-cause and CVD mortality, which is in line with our results.

The ‘obesity paradox’ would somehow contradict the concept of overweight and obesity being a strong risk factor for the development of cardiovascular diseases. Our results actually suggest that obesity with BMI > 30 kg/m^2^ is also a risk factor for increased mortality in AMI patients and in this sense consistent with the concept of obesity being a cardiovascular risk factor not only in primary (and secondary), but also in tertiary prevention.

### Low BMI values and long-term mortality

In prior publications it has been reported, that lower BMI < 25 kg/m^2^ is associated with increased long-term mortality in CAD and AMI ^7,8,11,12,21,22^, which is well in line with the results of the present study. However, there are studies reporting contradicting results ^16^. In the above mentioned study by Al-Shaar et al. for instance, lower BMI values were only non-significantly associated with slightly increased mortality ^16^. They speculated, that this might be an artifact caused by unintentional weight loss due to preexisting disease (reverse causation). Especially, patients with preexisting cancer might drive this potential reverse causation, whichalso might be applicable to the present study. However, our results were very clear in this respect: we observed a higher mortality for lower BMI not only for all-cause and cancer mortality, but also for CVD mortality and IHD mortality. Moreover, we performed a sensitivity analysis for cancer mortality by excluding all patients with an observational time of less than 2 years, thus excluding patients with preexisting, severe and advanced underlying diseases such as cancer. But still, BMI values < 25 kg/m^2^ were strongly associated with higher mortality. Therefore, the here reported associations between lower BMI and higher mortality are regarded as valid and important findings.

### All-cause mortality and age groups

While there was a pronounced U-shaped association between BMI and all-cause mortality for AMI patients younger than 60 years, there was no association between higher BMI values and mortality in the elderly group. We can only speculate about the reason for this observation: Younger AMI patients have a higher life expectancy after their infarction compared to older patients. Prior studies reported that a longer duration of obesity is associated with increased risk of developing secondary diseases like diabetes mellitus ^23,24^ or CDV ^25^. So, obesity might reveal more distinct impacts on health in younger patients, who live long enough to develop secondary conditions and are more likely to be affected by life-threatening complications. Older patients in their seventies and eighties on the other hand often struggle to keep their weight: Many are especially affected by sarcopenia, which impacts on bodily functions, including the immune response to infections ^26^. Thus, at older age, a higher BMI likely represent higher muscle mass and bigger energy reserves and consequently less frailty. This circumstance might annihilate the effect of obesity-related complications like diabetes and CVD and consequently leading to comparable mortality for obese AMI patients compared to individuals with BMI < 30 kg/m^2^.

### Mortality and smoking

Many prior studies indicated effects of residual confounding by smoking and/or an interaction between BMI and smoking in mortality analyses ^3,16,27–29^. This appears to be not much of a surprise as smoking suppresses appetite which consequently can result in a relevant weight reduction ^30^. At the same time, smoking causes premature mortality since cigarette smoke contains a variety of carcinogenic compounds ^31^ and is involved in the pathophysiology of arteriosclerosis ^32,33^. To reflect this circumstance, we adjusted all models for the smoking status of the patients. Moreover, we performed a sensitivity analyses only including patients who never smoked cigarettes. This had moderate impact on the associations observed, and the overall shape of the associations was quite similar. Although smoking might be an important factor or confounder regarding the association between BMI and long-term mortality, the results found in this study appear to be applicable mainly independent of the smoking status (even though the effect sizes (HR’s) were attenuated).

### Strengths and Limitations

The present study uses data from the population-based Myocardial Infarction Registry Augsburg with complete enrollment, which minimizes selection bias and ensure highest data quality. The number of included AMI cases is high and the follow up of patients is long with a median observational time of 6.7 years. The broad variety of information on comorbidities, treatment etc. enabled the calculation of most complete multivariable-adjusted COX regression models. A particular strength of this study is information on the main cause of death and separate analyses for specific types of mortality.

Nevertheless, there are also some limitations to mention. First, information on BMI was only available for the time of infarction, and there was no follow-up information on BMI. BMI might have changed considerable over the course of up to ten years. Indeed, body-weight fluctuations are quite common ^34^ and go along with increased mortality in CAD and AMI patients ^34,35^. Additionally, measures of body composition, like ‘percentage of body fat’ or ‘body fat distribution’ might provide important additional information for the analysis of obesity/body composition and mortality ^36,37^. Furthermore, over the course of 17 years, which is the time frame for patient recruitment for this analysis, diagnostics, acute treatment and prevention of coronary artery diseases and AMI has changed significantly (PCI treatment, troponin diagnostics, medication, etc.), which may have affected the observed associations as well. Our findings may not be generalized to all ethnic groups since no information on ethnicity was available. Finally, we may have not considered all important confounders and we cannot exclude reverse causation.

## 5. Conclusion

There is a general U-shaped association between BMI and all-cause mortality in AMI patients. This association was most dominant in AMI patients < 60 years; in the elderly group high BMI was not associated with increased mortality. For cancer mortality, only BMI values < 25 kg/m^2^ were associated with increased mortality; individuals with obesity (BMI > 30kg/m^2^) on the other hand were not more likely to die from cancer. We conclude that patients with a mid-range BMI between 25 and 30 kg/m^2^ have the most favorable outcome after AMI; especially very low BMI values < 22 kg/m^2^ are associated with higher mortality.

## Data Availability

The datasets generated during and/or analysed during the current study are not publicly available due to data protection aspects but are available in an anonymized form from the corresponding author on reasonable request.

## Acknowledgements

We would like to thank all members of the Helmholtz Zentrum München, Institute of Epidemiology, and the Chair of Epidemiology at the University Hospital of Augsburg, who were involved in the planning and conduct of the study. Steering partners of the MI Registry, Augsburg, include the Chair of Epidemiology at the University Hospital of Augsburg and the Department of Internal Medicine I, Cardiology, University Hospital of Augsburg. Many thanks for their support go to the local health departments, the office-based physicians and the clinicians of the hospitals within the study area. Finally, we express our appreciation to all study participants.

## Authorś contribution

TS and CM conceived the study. TS performed the statistical analysis and drafted the manuscript. CM was responsible for the acquisition of the data and supervised the analysis. DF, PR and JL contributed to data acquisition and revised the manuscript. All authors approved the final manuscript.

## Ethics approval and consent to participate

Data collection of the MONICA/KORA MI registry has been approved by the ethics committee of the Bavarian Medical Association (Bayerische Landesärztekammer) and the study was performed in accordance with the Declaration of Helsinki. All study participants have given written informed consent.

